# Association of Obstructive Sleep Apnea With Risk of Myocardial Infarction: A Multivariable Mendelian Randomization and Mediation Analysis

**DOI:** 10.64898/2026.03.29.26349673

**Authors:** Yifan Zhang, Xiaoli Zhu, Yulan Chen, Ayiguzaili Maimaitimin, Zeyu Liang, Rehanguli Maihemutijiang, Nayiman Nihimaiti

## Abstract

**BACKGROUND:** Although observational studies have frequently reported an association between obstructive sleep apnea (OSA) and myocardial infarction (MI), whether this relationship is causal or primarily attributable to shared risk factors remains uncertain.

**METHODS AND RESULTS:** We conducted a 2-sample Mendelian randomization (MR) study to evaluate the causal effect of OSA on MI. Summary statistics for OSA were obtained from FinnGen (8,998 cases and 356,128 controls),and summary statistics for MI were obtained from the UK Biobank (14,394 cases and 361,194 controls),with validation performed using data from the CARDIoGRAMplusC4D consortium. Mediation MR was further performed to assess 13 potential mediators, including obesity, glycemic traits, blood pressure, and lipid-related traits. In addition, a 6-step multivariable MR (MVMR) framework was used to estimate the direct effect of OSA after adjustment for different sets of potential confounders. Reverse MR analysis was also conducted to assess possible reverse causality. The MR analysis showed that genetically predicted OSA liability was significantly associated with an increased risk of MI (odds ratio [OR] per log-OR increase, 1.0024 [95% CI, 1.0010–1.0039]; P=0.001). Mediation analysis identified body mass index (BMI) as the most prominent mediator, with an explained proportion of 35.94% P=0.030, whereas systolic blood pressure (SBP) showed only minimal mediation 0.28%; P=0.678. In the stepwise MVMR analysis, the association between OSA and MI was markedly attenuated after simultaneous adjustment for BMI and SBP P=0.156, suggesting that the observed effect may partly reflect a shared clinical baseline. Notably, in a head-to-head model adjusting for SBP and atrial fibrillation (AF), AF remained a robust independent risk factor P=0.004, whereas OSA retained only a marginal direct effect P=0.050, indicating that the OSA–AF axis may operate independently of chronic pressure load. Reverse MR analysis found no evidence that genetic liability to MI influenced the risk of OSA.

**CONCLUSIONS:** Our study provides genetic evidence supporting a causal association between OSA and MI and suggests that this relationship may be mediated, in part, through obesity-related and arrhythmia-related pathways. In particular, AF may represent an important intermediate node linking OSA to MI, independent of traditional hemodynamic factors such as SBP, which may have implications for the clinical management of OSA.

## Introduction

Cardiovascular diseases remain the leading cause of disease burden and death worldwide, and myocardial infarction (MI) continues to account for a substantial proportion of cardiovascular mortality^1,2^. In addition to the well-recognized risk factors for MI, such as obesity, smoking, hypertension, diabetes, and dyslipidemia, accumulating observational studies have suggested that obstructive sleep apnea (OSA) is associated with an increased risk of MI^3^.

OSA, characterized by recurrent upper airway collapse during sleep, is a common chronic inflammatory disorder with a high prevalence in the adult population^4,5^.OSA is frequently accompanied by cardiovascular comorbidities, and previous observational studies have suggested that the association between OSA and MI may be partly mediated by environmental and clinical factors, particularly obesity and smoking^6^. However, because observational studies are susceptible to residual confounding and potential reverse causation, the true causal effect of OSA on MI remains uncertain^7^.

Under certain assumptions, Mendelian randomization (MR) can estimate the causal relationship between an exposure and an outcome by using genetic variants as instrumental variables, thereby strengthening causal inference by reducing bias from confounding and reverse causation^8^. Summary statistics from large-scale genome-wide association studies provide reliable instrumental variables for MR analyses. To date, however, genetic evidence on the relationship between obstructive sleep apnea and myocardial infarction remains limited^9^.

In the present study, we performed a 2-sample MR analysis to investigate the causal association between genetically predicted OSA and MI risk. We further used 2-step MR analyses and progressive multivariable MR to evaluate the mediating roles of obesity-related traits, metabolic factors, and cardiovascular or behavioral intermediate phenotypes and to assess the residual independent effect of OSA on MI. This study was designed to provide genetic epidemiological evidence for the complex causal network linking OSA and MI and to offer clues for targeted cardiovascular risk prevention.

## Methods

### Study Design

All data used in this 2-sample Mendelian randomization (MR) study were obtained from publicly available genome-wide association studies (GWASs), and detailed information on the data sources is provided below. We conducted the MR study to evaluate the causal association between genetically predicted obstructive sleep apnea (OSA) and myocardial infarction (MI). An overview of the bidirectional MR study design is shown in Figure 1. To minimize potential bias attributable to population stratification, all participants included in the analyses were restricted to individuals of European ancestry. This study was performed in accordance with the STROBE-MR guideline^10^. Because all materials were derived from publicly available anonymized summary-level datasets, no additional ethical approval was required.

**Figure 1.**
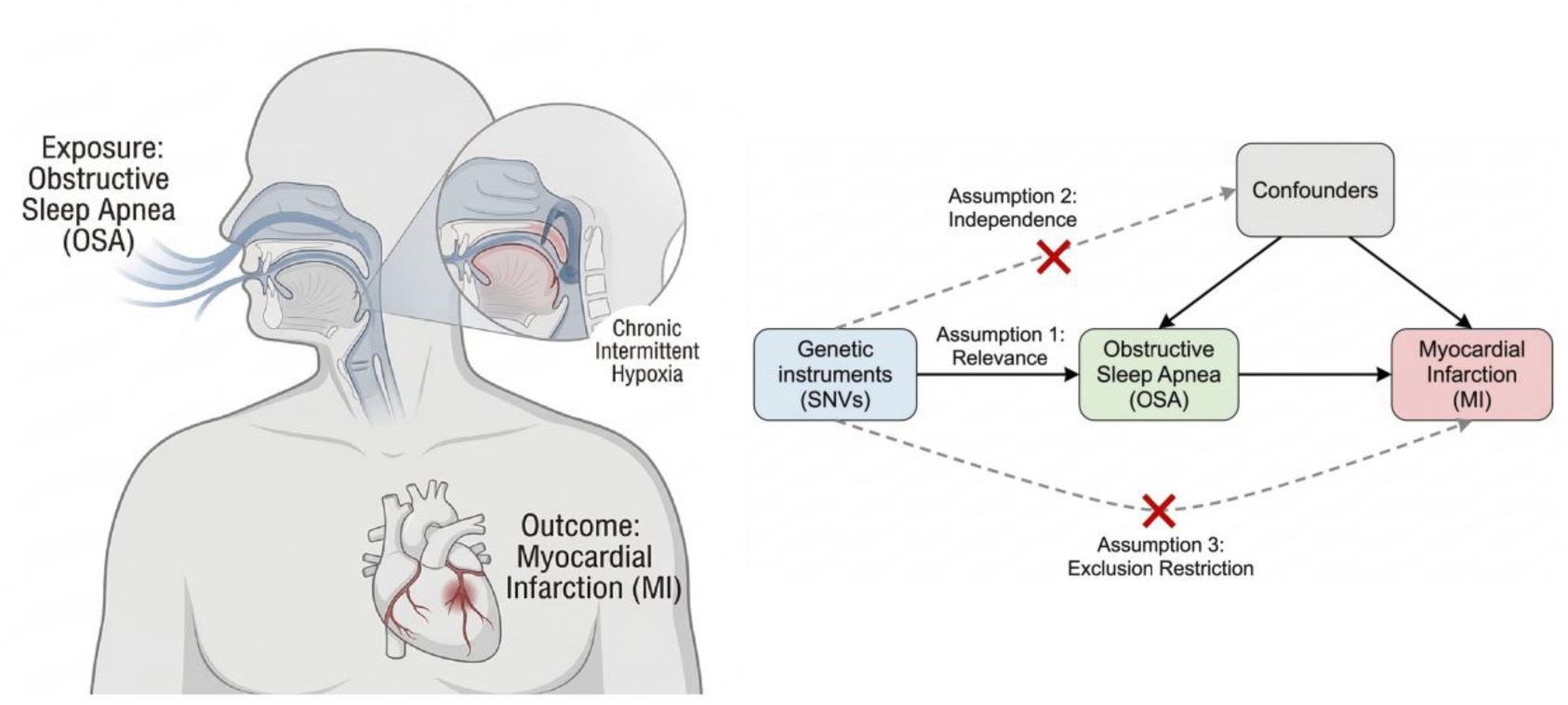
Conceptual framework and Mendelian randomization (MR) design of the study. The left panel illustrates the pathophysiological pathway from obstructive sleep apnea (OSA) to myocardial infarction (MI) via chronic intermittent hypoxia. The right panel displays the directed acyclic graph (DAG) of the MR analysis using genetic instruments (SNVs). The three core instrumental variable (IV) assumptions are indicated: (1) Relevance: genetic variants are robustly associated with OSA; (2) Independence: variants are not associated with potential confounders; (3) Exclusion restriction: variants affect MI risk only through OSA. Red crosses denote the violation of paths to ensure the validity of IV assumptions. Abbreviations: OSA, obstructive sleep apnea; MI, myocardial infarction; SNV, single nucleotide variant.

**Figure 2.**
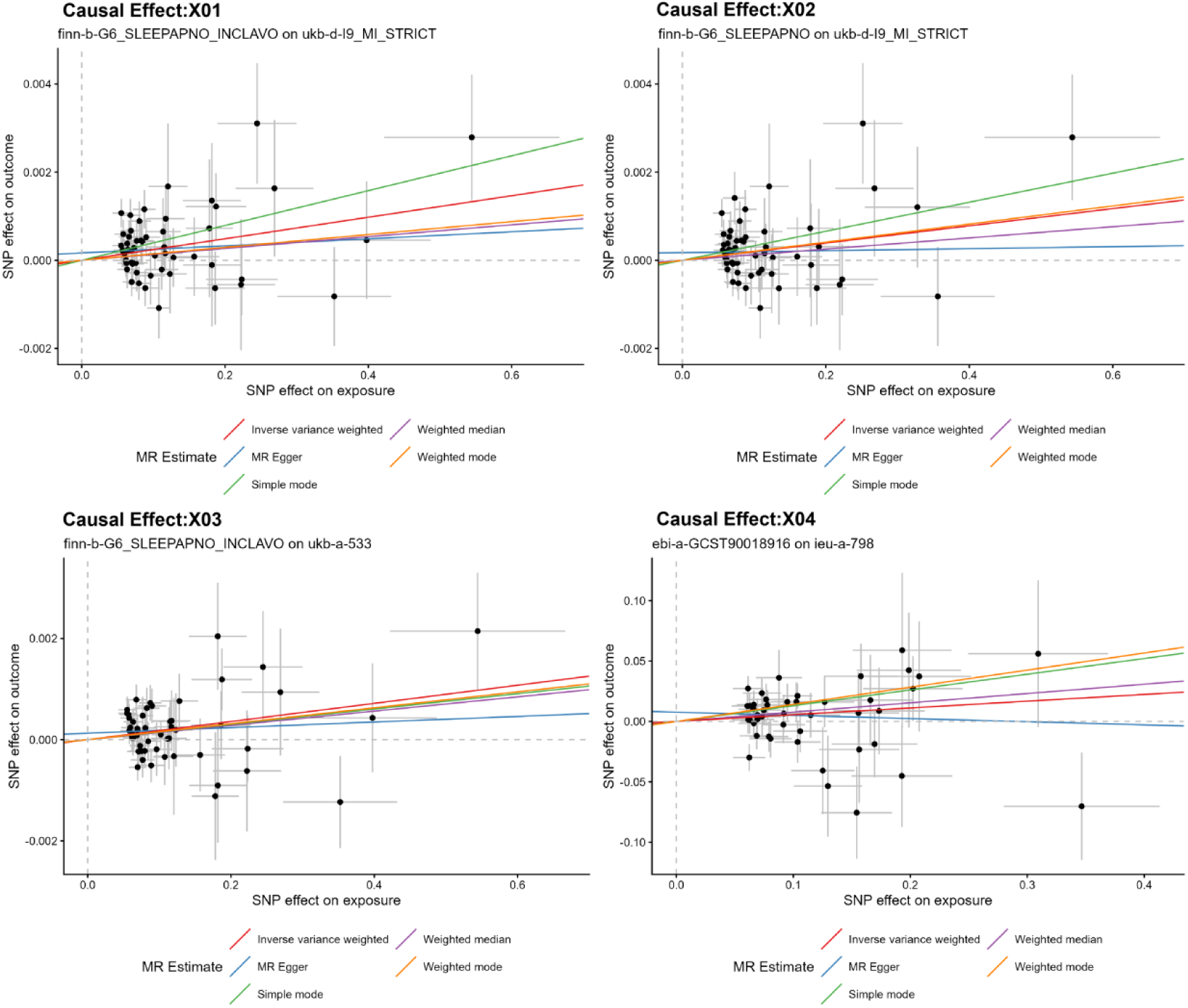
Scatter plots of SNV effects on OSA versus MI across four independent analysis groups. Each point represents a single nucleotide variant (SNV), with horizontal and vertical error bars indicating the standard error of the effect on OSA (exposure) and MI (outcome), respectively. The slopes of the colored lines correspond to the causal estimates derived from five Mendelian randomization (MR) methods: Inverse variance weighted (red), MR Egger (blue), Weighted median (purple), Weighted mode (orange), and Simple mode (green). Panels (A–D) correspond to different data source combinations (X01–X04), illustrating the consistency of the causal direction across datasets. Abbreviations: OSA, obstructive sleep apnea; MI, myocardial infarction; SNV, single nucleotide variant; IVW, inverse variance weighted.

### Data Sources and SNP Selection for OSA

Single-nucleotide polymorphisms (SNPs) were used as instrumental variables (IVs) in the MR analysis. Summary-level data for OSA in the primary analysis were obtained from FinnGen release 10(ID:finn-b-G6_SLEEPAPNO_INCLAVO), including 45,116 cases and 351,194 controls^11^. A series of screening procedures were conducted to select eligible IVs. First, SNPs associated with OSA at the genome-wide significance threshold of P<5×10^-8 were selected as candidate IVs; when necessary, the threshold was relaxed to P<1×10^-5 to improve instrument availability. Sensitivity analyses were conducted to ensure that the inclusion of suggestive SNPs did not bias the primary estimates. Second, linkage disequilibrium clumping (r^2<0.001, clump window=10,000 kb) was performed to retain independent SNPs. Third, SNP-specific F statistics were calculated, and only SNPs with F statistics >10 were retained to reduce weak instrument bias. Fourth, the proportion of variance explained (R^2^) by the selected instruments was estimated using the effect allele frequency and the corresponding genetic effect estimates. Fifth, Steiger filtering was applied to exclude SNPs showing evidence of reverse directionality. Finally, to further reduce potential pleiotropic bias, the selected SNPs were systematically queried in the LDtrait module of the LDlink database, and variants associated at genome-wide significance with known cardiometabolic confounders were flagged and removed in sensitivity analyses.

### Data Sources for MI

Summary-level data for MI in the primary analysis were obtained from the UK Biobank (ID:ukb-d-I9_MI_STRICT), including 7,018 cases and 354,176 controls^12^. MI was defined according to strict clinical criteria in the original GWAS.

### Validation Analyses

To assess the robustness and generalizability of the findings, we further performed 3 validation analyses. First, for sensitivity validation of the exposure phenotype, we used an alternative OSA dataset from FinnGen based only on standard inpatient diagnoses(ID:finn-b-G6_SLEEPAPNO), including 38,710 cases and 351,194 controls^11^. Second, for sensitivity validation of the outcome phenotype, we used acute MI summary statistics from the UK Biobank based on a different diagnostic definition(ID:ukb-a-533), including 3,927 cases and 333,272 controls^12^. Third, for independent cross-cohort validation, we introduced a fully independent OSA dataset(ID:ebi-a-GCST90018916), including 13,818 cases and 463,035 controls^13^,together with clinically confirmed MI summary statistics from the CARDIoGRAMplusC4D consortium(ID:ieu-a-798), including 43,676 cases and 128,199 controls^14^.

### Statistical Analysis

Inverse-variance weighting (IVW) with a random-effects model was used as the primary method to estimate the causal effect of OSA on MI. Weighted median analysis and MR-Egger regression were additionally performed as complementary methods. To evaluate the robustness of the findings, we assessed directional horizontal pleiotropy using the MR-Egger intercept, detected and corrected outliers using the MR-PRESSO global test, and performed leave-one-out analyses to evaluate whether any single SNP disproportionately influenced the overall causal estimate. Because partial sample overlap may exist in UK Biobank-based analyses, we additionally assessed its potential influence on type I error inflation.

### Two-step MR Mediation Analysis

To quantify potential mediation pathways, we performed 2-step MR analyses including 13 metabolic and cardiovascular phenotypes that were independent of the outcome cohort as candidate mediators. Indirect effects were estimated using the product of coefficients method, and standard errors were calculated using the Delta method. Mediation proportions were then estimated for each candidate pathway.

### Multivariable MR Analysis

To further evaluate the residual independent effect of OSA on MI after accounting for correlated risk factors, we conducted progressive multivariable MR (MVMR) analyses. In model 1, OSA and body mass index (BMI) were included to establish a basic adiposity-adjusted framework. Model 2 further included systolic blood pressure (SBP). Model 3 additionally incorporated waist-to-hip ratio (WHR) to distinguish general obesity from central adiposity. Model 4 further adjusted for the metabolic network, including type 2 diabetes, low-density lipoprotein cholesterol, and triglycerides. Model 5 additionally included smoking status and atrial fibrillation (AF). In model 6, a head-to-head adjustment for AF and SBP was performed to compare the independent contribution of electrophysiological remodeling and hemodynamic load to MI risk. In MVMR analyses, instrument strength was evaluated using the Sanderson-Windmeijer conditional F statistic, and a conditional F statistic >10 was considered indicative of adequate instrument strength.

### Software and Statistical Thresholds

All statistical analyses were performed using R version 4.5.1, primarily with the TwoSampleMR package. A 2-sided P<0.05 was considered statistically significant, and false discovery rate correction was applied for multiple testing where appropriate.

## RESULTS

Analysis SetsAfter evaluation of phenotype consistency and quality control for sample overlap, X01, defined as extended-definition obstructive sleep apnea (OSA) from FinnGen and strictly defined myocardial infarction (MI) from the UK Biobank, was selected as the primary analysis set. X02, X03, and X04 were further used as validation sets to assess the consistency of the findings across different phenotype definitions and study platforms.

### Characteristics of the Selected SNPs

At the conventional genome-wide significance threshold (P<5×10^−8^, only 5 independent single-nucleotide polymorphisms (SNPs) were identified as instrumental variables (IVs) for OSA. Although this set showed a high mean F statistic (40.33), the explained variance was limited, and the IVW analysis did not provide sufficient evidence for an association between genetically predicted OSA and MI (odds ratio [OR], 1.001 [95% CI, 0.993–1.008]). In addition, the small number of IVs did not allow reliable pleiotropy testing or subsequent multivariable Mendelian randomization (MVMR) analyses.

Therefore, to balance statistical power and the feasibility of sensitivity analyses, we selected SNPs at P<1×10^-5 for the primary analyses. Under this criterion, 54 independent SNPs were identified, explaining approximately 0.33% of the phenotypic variance in OSA. The mean F statistic was 23.19, indicating a low likelihood of weak instrument bias. Based on this instrument set, the IVW analysis showed that genetically predicted OSA was significantly associated with an increased risk of MI (OR, 1.002 [95% CI, 1.001–1.004]; P=0.001)(Table 1,Figure2).

**Table 1.** Mendelian randomization estimates of the causal effect of obstructive sleep apnea on myocardial infarction risk across different datasets.

| Model ID | Exposure Source | Outcome Source | No. of IVs | Mean F-statistic | OR (95% CI) | P-value (IVW) | Heterogeneity P-value (Q-test) | Pleiotropy P-value (MR-Egger) |
| --- | --- | --- | --- | --- | --- | --- | --- | --- |
| X01 | FinnGen (Incl. Avohilmo) | UK Biobank (Strict MI) | 54 | 23.19 | 1.002 (1.001 – 1.004) | 0.001 | 0.141 | 0.273 |
| X02 | FinnGen (Standard Inpatient) | UK Biobank (Strict MI) | 55 | 23.12 | 1.002 (1.001 – 1.004) | 0.008 | 0.182 | 0.259 |
| X03 | FinnGen (Incl. Avohilmo) | UK Biobank (Acute MI) | 53 | 23.21 | 1.002 (1.001 – 1.004) | 0.003 | 0.236 | 0.291 |
| X04 | EBI (ebi-a-GCST90018916) | CARDIoGRAMplusC4D | 46 | 23.24 | 1.057 (1.000 – 1.118) | 0.05 | 0.158 | 0.279 |

Notes: OR, odds ratio; CI, confidence interval; IVW, inverse variance weighted; IV, instrumental variable; OSA, obstructive sleep apnea; MI, myocardial infarction.
Data Sources: "Strict MI" and "Acute MI" definitions in UK Biobank were based on ICD-10 codes. Instrument Strength: A mean F-statistic $> 10$ was used to indicate the absence of weak instrument bias. Methodology: The primary results are based on the IVW method.

Overall, the direction of the effect estimates was consistent across different SNP selection thresholds, and all instrument sets showed adequate strength on the basis of F statistics. Therefore, all subsequent primary analyses, sensitivity analyses, and multivariable analyses were based on the SNP set selected at P<1×10^-5. Under the observed variance explained(R^2^=0.33%), significance level α=0.05, and outcome sample size (457,824 participants, including 10,951 cases), the statistical power of the primary analysis exceeded 89%.

### Causal Effects of OSA on MI

The primary MR analysis showed that genetically predicted OSA was significantly associated with an increased risk of MI in the IVW analysis (OR, 1.002 [95% CI, 1.001–1.004]; P=0.001). No significant heterogeneity was detected in the Cochran Q test P=0.141, and the MR-Egger intercept did not indicate directional horizontal pleiotropy P=0.273. In addition, the MR-PRESSO global test did not identify significant outliers, suggesting that the primary model was robust.

After querying all candidate IVs in the LDtrait database, 21 variants associated with known cardiometabolic confounding phenotypes were removed. Using the remaining 36 IVs (mean F statistic, 22.00), the IVW analysis still showed a statistically significant association between OSA and MI (OR, 1.002 [95% CI, 1.000–1.004]; P=0.025). No significant heterogeneity was observed in the Cochran Q test P=0.853, no directional pleiotropy was detected in the MR-Egger intercept analysis P =0.987, and no significant outliers were identified in the MR-PRESSO global test P=0.849. Although the weighted median estimate did not reach statistical significance P=0.227, the direction of effect was consistent with the IVW analysis. Steiger directionality testing supported the causal direction from OSA to MI (P<0.001), and leave-one-out analysis showed that the ORs ranged from 1.0017 to 1.0021 after sequential removal of individual SNPs, suggesting that the observed association was not driven by any single variant.

### Causal Effects of MI on OSA

Reverse-direction MR analysis was performed to investigate whether genetic liability to MI influenced the risk of OSA. Using 9 IVs with strong instrument strength (mean F statistic, 63.03), we found no evidence that genetically predicted MI affected OSA risk in the IVW analysis (OR, 0.26 [95% CI, 0.02–3.67]; P=0.482). No significant heterogeneity P=0.330 or directional horizontal pleiotropy P=0.053 was detected. In addition, the MR Steiger test supported the correct causal direction in all analysis sets (X01–X04; Steiger proportion correct=1). Taken together, these findings provided no evidence for reverse causality from MI to OSA.

### Sensitivity and Validation Analyses

To assess the robustness and external validity of the primary findings, we performed a series of validation analyses using alternative phenotype definitions and independent cohorts. In analyses of strictly defined MI in the UK Biobank, significant risk effects were observed when either the inpatient-diagnosed OSA dataset or the extended OSA dataset was used as the exposure (P<0.05 for both comparisons). When the outcome was replaced by acute MI defined according to a different diagnostic standard, the positive causal association was also replicated (OR, 1.002 [95% CI, 1.001–1.003]; P=0.003). In addition, independent cross-cohort validation using an external OSA dataset and clinically confirmed MI summary statistics from the CARDIoGRAMplusC4D consortium yielded a consistent significant result (OR, 1.057 [95% CI, 1.000–1.118]; P=0.0499). Although the point estimates varied slightly across datasets, the direction of effect remained consistent, supporting OSA as a stable genetic risk factor for MI (Table 1).

Additional sensitivity analyses further supported the robustness of the findings. The MR-Egger intercept remained close to the null (P>0.05), the Cochran Q test did not indicate significant heterogeneity (P>0.05), and the regression slopes from the IVW, weighted median, and weighted mode methods were concordant. Leave-one-out analysis also confirmed that the causal estimate was not disproportionately driven by any single SNP.

### Two-step MR Mediation Analyses

We next performed 2-step MR analyses to evaluate the potential mediating roles of 13 metabolic and cardiovascular-related traits in the pathway linking OSA to MI. Among all candidate mediators, only body mass index (BMI) showed a statistically significant indirect effect. The estimated indirect effect mediated through BMI was 0.00088 (95%CI,8.63×10^−5^ to 0.00167; P=0.030 false discovery rate [FDR]-adjusted P=0.386), corresponding to a mediation proportion of 35.94% (95% CI, 3.54%–68.35%). In contrast, type 2 diabetes P=0.152, low-density lipoprotein cholesterol and triglycerides both P=0.487, uric acid P=0.282, systolic blood pressure (SBP; P=0.678; mediation proportion, 0.28%), and the remaining candidate factors did not show statistically significant mediation effects(Table 2,Figure 3).

**Figure 3.**
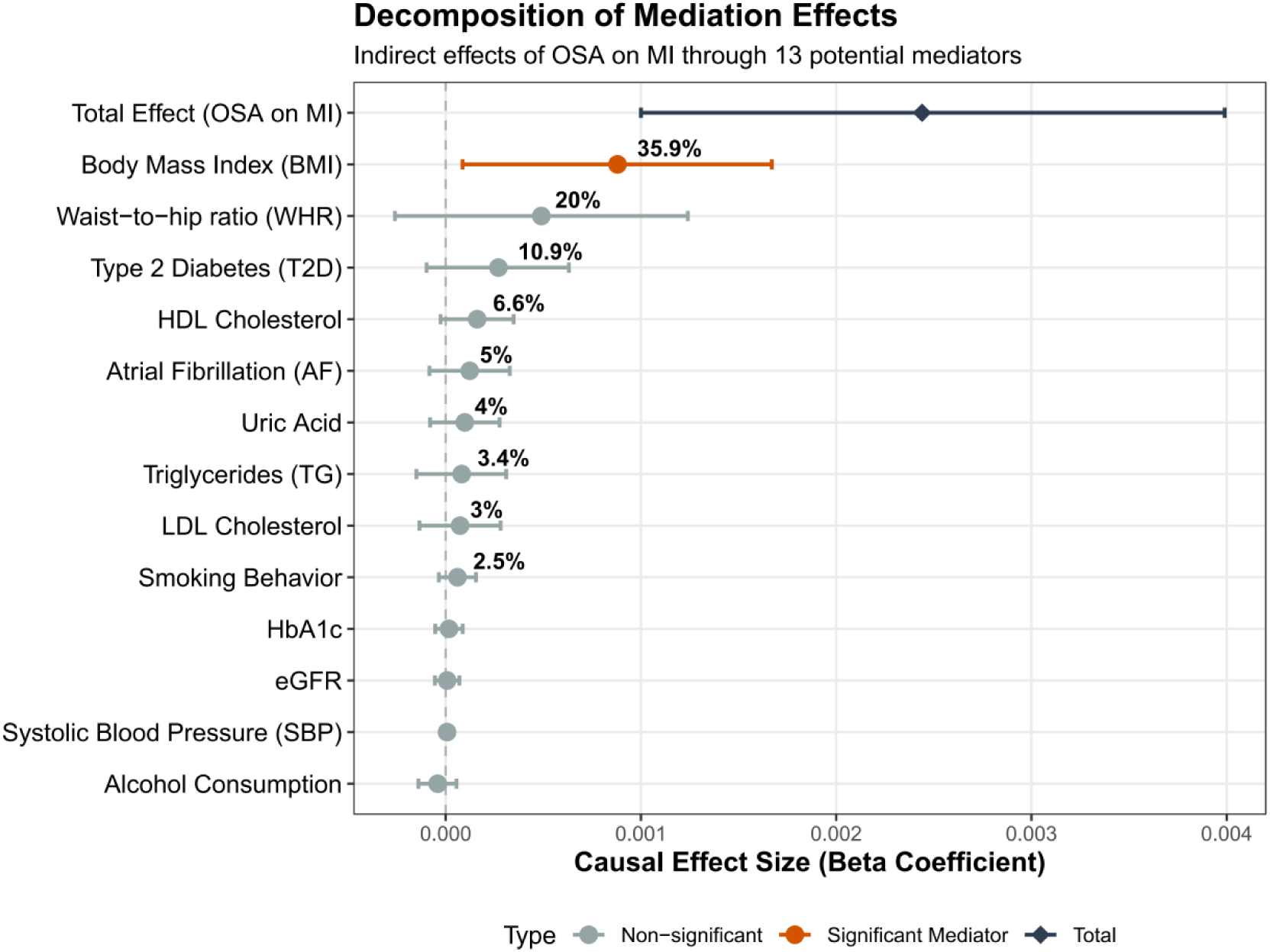
Mediation analysis of the causal effect of OSA on MI risk via 13 potential metabolic and lifestyle mediators. The “Total Effect” (top) represents the overall causal impact of OSA on MI. For each potential mediator, the horizontal bars indicate the indirect effect size (beta coefficient) with 95% confidence intervals (CIs). The percentages (e.g., 35.9% for BMI) denote the Proportion Mediated (PM), calculated as (Indirect Effect / Total Effect). Orange markers indicate mediators with statistically significant indirect effects (P < 0.05), while grey markers represent non-significant paths. Abbreviations: BMI, body mass index; WHR, waist-to-hip ratio; T2D, type 2 diabetes; AF, atrial fibrillation; TG, triglycerides; LDL, low-density lipoprotein; HbA1c, glycated hemoglobin; eGFR, estimated glomerular filtration rate; SBP, systolic blood pressure.

**Table 2.**
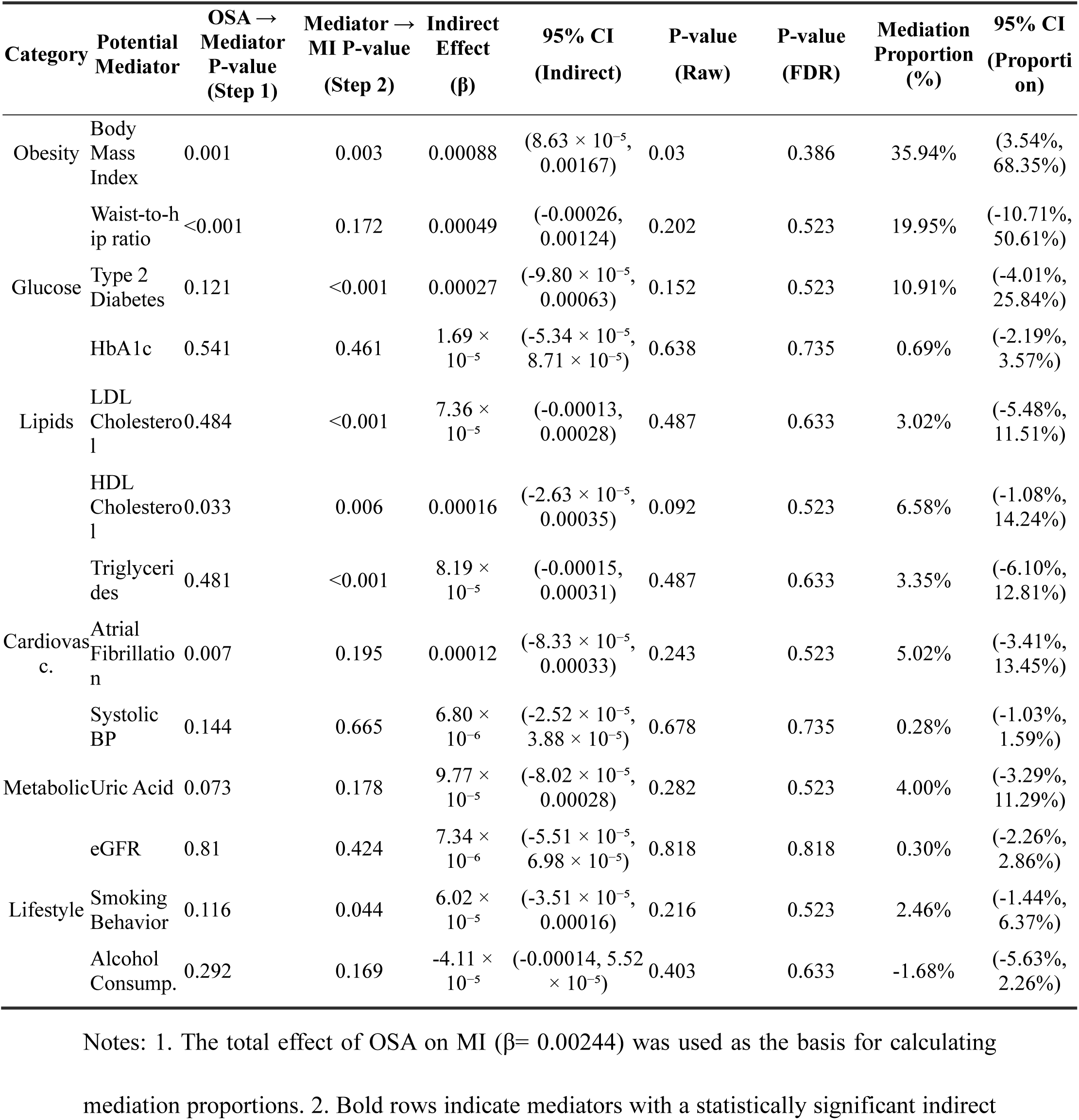
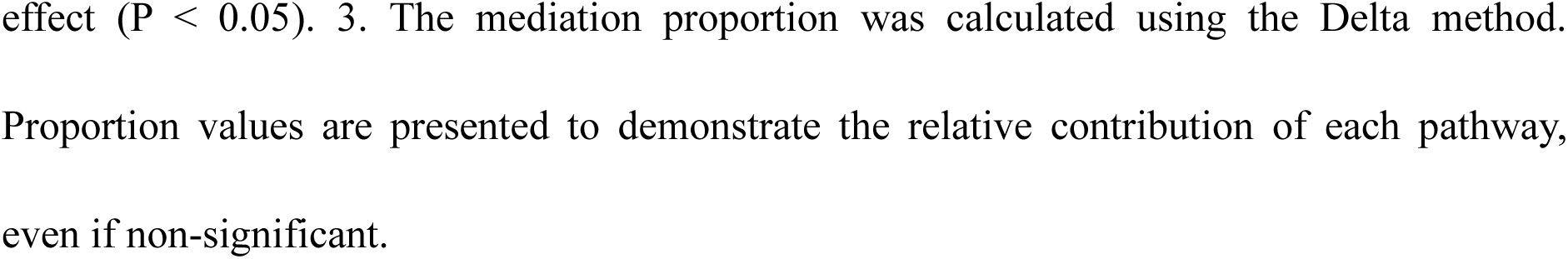
Mediation analysis of the causal effect of OSA on MI risk through metabolic and cardiovascular factors.

| Category | Potential Mediator | OSA → Mediator P-value (Step 1) | Mediator → MI P-value (Step 2) | Indirect Effect (β) | 95% CI (Indirect) | P-value (Raw) | P-value (FDR) | Mediation Proportion (%) | 95% CI (Proportion) |
| --- | --- | --- | --- | --- | --- | --- | --- | --- | --- |
| Obesity | Body Mass Index | 0.001 | 0.003 | 0.00088 | $(8.63 \times 10^{-5}, 0.00167)$ | 0.03 | 0.386 | 35.94% | (3.54%, 68.35%) |
| | Waist-to-hip ratio | <0.001 | 0.172 | 0.00049 | $(-0.00026, 0.00124)$ | 0.202 | 0.523 | 19.95% | (-10.71%, 50.61%) |
| Glucose | Type 2 Diabetes | 0.121 | <0.001 | 0.00027 | $(-9.80 \times 10^{-5}, 0.00063)$ | 0.152 | 0.523 | 10.91% | (-4.01%, 25.84%) |
| | HbA1c | 0.541 | 0.461 | $1.69 \times 10^{-5}$ | $(-5.34 \times 10^{-5}, 8.71 \times 10^{-5})$ | 0.638 | 0.735 | 0.69% | (-2.19%, 3.57%) |
| Lipids | LDL Cholesterol | 0.484 | <0.001 | $7.36 \times 10^{-5}$ | $(-0.00013, 0.00028)$ | 0.487 | 0.633 | 3.02% | (-5.48%, 11.51%) |
| | HDL Cholesterol | 0.033 | 0.006 | 0.00016 | $(-2.63 \times 10^{-5}, 0.00035)$ | 0.092 | 0.523 | 6.58% | (-1.08%, 14.24%) |
| | Triglycerides | 0.481 | <0.001 | $8.19 \times 10^{-5}$ | $(-0.00015, 0.00031)$ | 0.487 | 0.633 | 3.35% | (-6.10%, 12.81%) |
| Cardiovascular | Atrial Fibrillation | 0.007 | 0.195 | 0.00012 | $(-8.33 \times 10^{-5}, 0.00033)$ | 0.243 | 0.523 | 5.02% | (-3.41%, 13.45%) |
| | Systolic BP | 0.144 | 0.665 | $6.80 \times 10^{-6}$ | $(-2.52 \times 10^{-5}, 3.88 \times 10^{-5})$ | 0.678 | 0.735 | 0.28% | (-1.03%, 1.59%) |
| Metabolic | Uric Acid | 0.073 | 0.178 | $9.77 \times 10^{-5}$ | $(-8.02 \times 10^{-5}, 0.00028)$ | 0.282 | 0.523 | 4.00% | (-3.29%, 11.29%) |
| | eGFR | 0.81 | 0.424 | $7.34 \times 10^{-6}$ | $(-5.51 \times 10^{-5}, 6.98 \times 10^{-5})$ | 0.818 | 0.818 | 0.30% | (-2.26%, 2.86%) |
| Lifestyle | Smoking Behavior | 0.116 | 0.044 | $6.02 \times 10^{-5}$ | $(-3.51 \times 10^{-5}, 0.00016)$ | 0.216 | 0.523 | 2.46% | (-1.44%, 6.37%) |
| | Alcohol Consump. | 0.292 | 0.169 | $-4.11 \times 10^{-5}$ | $(-0.00014, 5.52 \times 10^{-5})$ | 0.403 | 0.633 | -1.68% | (-5.63%, 2.26%) |

Notes: 1. The total effect of OSA on MI ( $\beta = 0.00244$ ) was used as the basis for calculating mediation proportions. 2. Bold rows indicate mediators with a statistically significant indirect
effect ( $P < 0.05$ ). 3. The mediation proportion was calculated using the Delta method. Proportion values are presented to demonstrate the relative contribution of each pathway, even if non-significant.

Reverse MR analysis further showed a strong positive causal effect of BMI on OSA (P<0.001), suggesting a bidirectional genetic relationship between these 2 traits. In addition, atrial fibrillation (AF) showed a more stable downstream pattern, with a stronger effect of OSA on AF P=0.007 than the reverse-direction signal P=0.021, supporting AF as a downstream pathological node in the OSA-related disease pathway. However, although the indirect effect of BMI reached nominal significance, it did not remain significant after false discovery rate correction for the 13 candidate mediators.

### Progressive Multivariable MR Analyses

We further constructed 6 progressive MVMR models to evaluate the residual direct effect of OSA on MI after adjustment for correlated risk factors. In model 1, adjustment for BMI attenuated the direct effect of OSA, although the association remained statistically significant (β=0.00195; P=0.038). In model 2, to distinguish general obesity from central adiposity, we adjusted for BMI and waist-to-hip ratio (WHR); the independent effect of BMI was abolished (P=0.652), whereas WHR emerged as a significant independent risk factor (P=0.017), and the direct effect of OSA remained significant (P=0.045). In model 3, after simultaneous adjustment for BMI and systolic blood pressure (SBP), SBP showed a strong independent effect (P<0.001), whereas the independent effects of OSA (P=0.156) and BMI (P=0.063) were no longer statistically significant.

In model 4, after adjustment for the metabolic network (type 2 diabetes, low-density lipoprotein cholesterol, and triglycerides), all metabolic traits showed strong effects (P values < 0.001), whereas OSA still retained a significant direct effect on MI (P=0.015). In model 5, adjusting for smoking and atrial fibrillation (AF) showed that both factors had significant independent effects, and the direct effect of OSA became marginal (P=0.064). Finally, in model 6, a head-to-head adjustment for AF and SBP was performed; AF showed a significant independent effect (P=0.004) despite the strong influence of SBP (P<0.001), whereas OSA retained a borderline direct effect (P=0.050) (Table 3, Figure 4).

**Figure 4.**
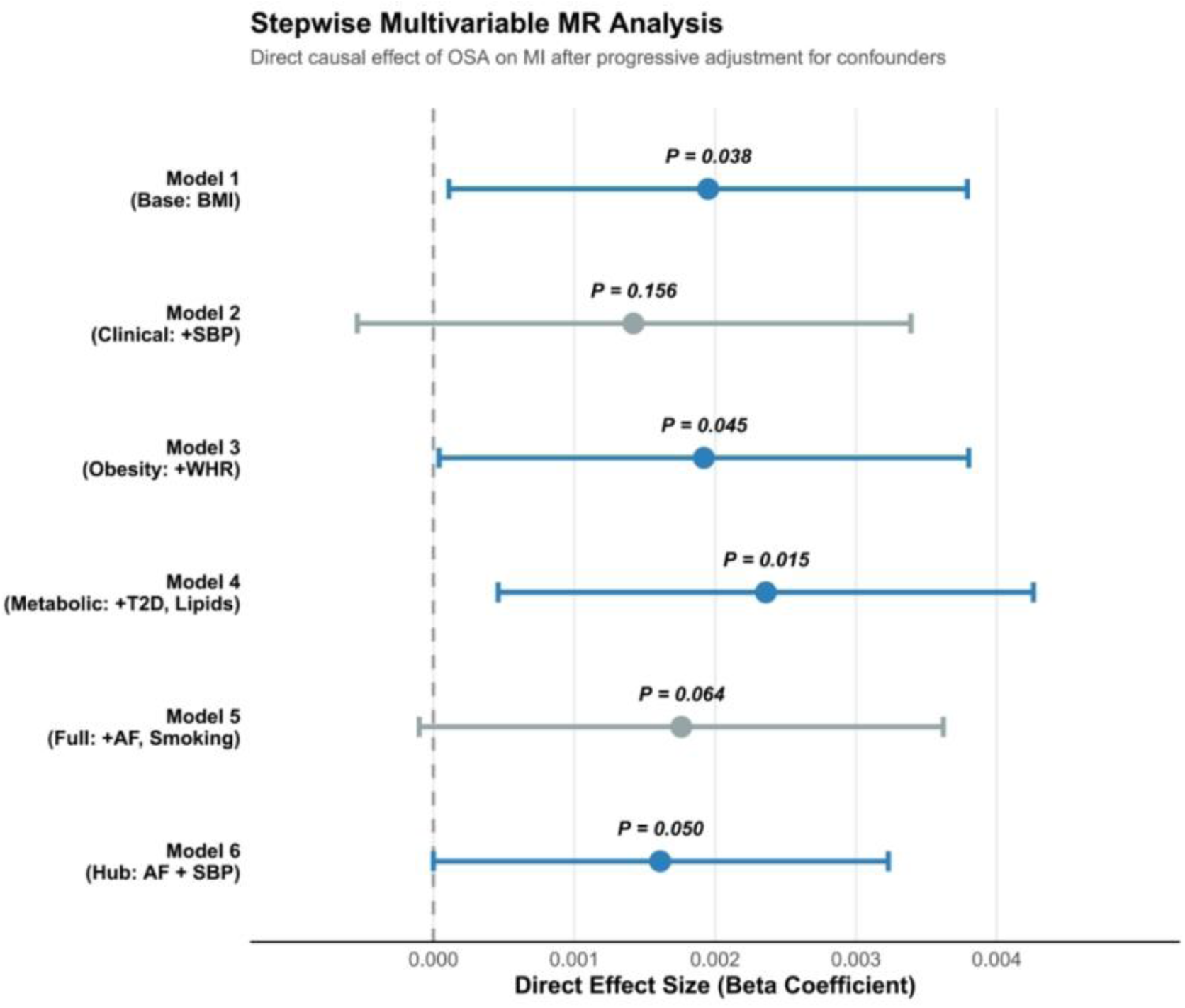
Multivariable Mendelian randomization (MVMR) estimates for the direct effect of OSA on MI risk. This forest plot presents the direct causal effect sizes (beta coefficients) of OSA on MI after progressive adjustment for potential pleiotropic pathways and mediators across six models. Model 1 serves as the baseline (adjusted for BMI). Subsequent models (2–6) incorporate clinical, metabolic, and lifestyle factors as indicated. Blue bars represent statistically significant direct effects (P ≤ 0.05), while grey bars denote non-significant associations. Horizontal error bars indicate 95% confidence intervals (CIs). Abbreviations: AF, atrial fibrillation; BMI, body mass index; MI, myocardial infarction; OSA, obstructive sleep apnea; SBP, systolic blood pressure; T2D, type 2 diabetes; WHR, waist-to-hip ratio.

**Table 3.** Direct causal effects of OSA on MI estimated by multivariable Mendelian randomization models.

| Model & Adjusted Factors | Direct Effect $\beta$ | 95% CI $\beta$ | P-value | SW F-statistic |
| --- | --- | --- | --- | --- |
| Model 1: Core Obesity (n=80) |  |  |  |  |
| Obstructive sleep apnea (OSA) | 0.00195 | (0.00011, 0.00379) | 0.038 | 52555.6 |
| Body mass index (BMI) | 0.00471 | (0.00118, 0.00823) | 0.009 | 87420.3 |
| Model 2: Obesity Dimensions (n=107) |  |  |  |  |
| Obstructive sleep apnea (OSA) | 0.00192 | (0.00004, 0.00379) | 0.045 | 25654 |
| Body mass index (BMI) | 0.00101 | (-0.00337, 0.00538) | 0.652 | 138770.6 |
| Waist-to-hip ratio (WHR) | 0.0076 | (0.00136, 0.01385) | 0.017 | 74491.4 |
| Model 3: Clinical Baseline Plus (New, n=148) |  |  |  |  |
| Obstructive sleep apnea (OSA) | 0.00142 | (-0.00054, 0.00339) | 0.156 | 4498.2 |
| Body mass index (BMI) | 0.00404 | (-0.00022, 0.00830) | 0.063 | 27500.8 |
| Systolic blood pressure (SBP) | 0.0158 | (0.01288, 0.01873) | < 0.001 | 52914.1 |
| Model 4: Metabolic Network (n=227) |  |  |  |  |
| Obstructive sleep apnea (OSA) | 0.00236 | (0.00045, 0.00426) | 0.015 | 11083.5 |
| Type 2 diabetes | 0.00273 | (0.00149, 0.00397) | < 0.001 | 497.5 |
| LDL cholesterol | 0.00747 | (0.00495, 0.00999) | < 0.001 | 7538.9 |
| Triglycerides | 0.00515 | (0.00256, 0.00773) | < 0.001 | 8115.4 |
| Model 5: Behavioral & Structural (n=116) |  |  |  |  |
| Obstructive sleep apnea (OSA) | 0.00176 | (-0.00010, 0.00363) | 0.064 | 18946.2 |
| Atrial fibrillation (AF) | 0.00141 | (0.00021, 0.00261) | 0.022 | 2176.7 |
| Cigarettes per day (Smoking) | 0.00328 | (0.00041, 0.00615) | 0.025 | 36340.5 |
| Model 6: Hub vs. Hemodynamics (New, n=124) |  |  |  |  |
| Obstructive sleep apnea (OSA) | 0.00161 | (0.00000, 0.00323) | 0.050 | 20174.4 |
| Atrial fibrillation (AF) | 0.00177 | (0.00058,<br>0.00297) | 0.004 | 1294.4 |
| Systolic blood pressure (SBP) | 0.01316 | (0.01030,<br>0.01603) | < 0.001 | 34090 |

Notes: AF, atrial fibrillation; BMI, body mass index; CI, confidence interval; LDL, low-density lipoprotein; MVMR, multivariable Mendelian randomization; OSA, obstructive sleep apnea; SBP, systolic blood pressure; SW F-statistic, Sanderson-Windmeijer F-statistic; WHR, waist-to-hip ratio. Direct Effect ( $\beta$ ): Represents the estimated direct causal effect of the exposure on MI risk per [unit] increase in OSA liability, independent of the adjusted factors in each model. Instrument Strength: The SW F-statistic was used to assess the strength of instrumental variables in the multivariable setting; a value $> 10$ typically indicates sufficient instrument strength. Sample Size (n): Refers to the number of independent genetic variants used as instrumental variables in each multivariable model.

Across all MVMR models, the Sanderson-Windmeijer conditional F statistics ranged from 497.5 to 138,770.6, indicating that weak instrument bias was unlikely. In addition, multivariable MR-Egger intercept analyses showed no evidence of significant directional horizontal pleiotropy across the models (P range, 0.645–0.920). Overall, these findings suggested that the effect of OSA on MI may be mediated through a multidimensional network involving adiposity, metabolic dysregulation, and secondary electrophysiological remodeling, further studies are needed to clarify the relative contributions of these pathways.

## DISCUSSION

Our study used large-scale genome-wide association study data and a Mendelian randomization framework to investigate the relationship between obstructive sleep apnea (OSA) and myocardial infarction (MI). The results supported a positive causal association between genetically predicted OSA and MI. In addition, 2-step Mendelian randomization suggested that body mass index (BMI) may partly mediate this association, whereas progressive multivariable Mendelian randomization analyses indicated that central adiposity and atrial fibrillation (AF) may represent more important downstream components of the risk pathway. By contrast, systolic blood pressure (SBP), although strongly associated with MI^15^, did not appear to be the dominant mediator in the OSA–MI pathway. Overall, these findings suggest that the effect of OSA on MI may involve multiple interconnected pathways rather than a single linear mechanism, and further studies are needed to clarify the underlying biology.

First, the primary 2-sample Mendelian randomization analysis showed that genetically predicted OSA was associated with an increased risk of MI. This finding is generally consistent with previous large observational and prospective studies showing that OSA is associated with higher cardiovascular risk^16,17^. From a genetic epidemiological perspective, our results support the hypothesis that OSA may act as an upstream risk factor for ischemic heart disease. However, the magnitude of effect was modest, which may not be unexpected. OSA is a clinically and biologically heterogeneous phenotype, and MI represents a relatively distal cardiovascular endpoint influenced by multiple intermediate processes; therefore, the effect of OSA on MI may be partly diluted when estimated within an MR framework^18,19^. Accordingly, a modest causal estimate does not necessarily imply limited etiological relevance, and the biological pathways through which OSA contributes to MI require further investigation.

The main association remained directionally consistent across sensitivity analyses, which supported the robustness of the primary finding. Although the instrument selection threshold for OSA was less stringent than the conventional genome-wide significance level, the selected instruments showed adequate strength, with mean F statistics well above the conventional threshold, and the primary analysis retained sufficient statistical power. However, the use of a less stringent threshold may still have increased the likelihood of including variants with less specific biological relevance to OSA, even if substantial weak instrument bias was unlikely. In addition, the absence of material evidence of directional pleiotropy and the consistency of effect direction across models further increased confidence in the causal estimate. Nevertheless, these observations do not fully exclude more subtle forms of horizontal pleiotropy, which may be difficult to detect for a complex trait such as OSA. These findings should therefore still be interpreted with appropriate caution, because OSA is a highly polygenic and clinically heterogeneous phenotype, and currently available genetic instruments may reflect general liability to OSA rather than a single uniform biological process^20,21^.

Among the candidate mediators, BMI showed the only nominally significant indirect effect, accounting for approximately 35.94% of the observed pathway from OSA to MI. This finding is broadly consistent with previous studies and current clinical understanding that adiposity plays an important role in OSA-related cardiovascular risk^22^. However, because adiposity is closely linked to OSA both biologically and epidemiologically, the estimated mediation proportion should not be interpreted as a precise partition of effect, but rather as a signal that adiposity may contribute to the observed association. Accordingly, the finding may be viewed as being consistent with the potential relevance of weight management in patients with OSA^23^. although it does not directly demonstrate that weight reduction would necessarily attenuate MI risk through this pathway. However, the mediation effect of BMI did not remain statistically significant after false discovery rate correction. Given that indirect effects are often modest in magnitude, this finding provides limited evidential strength and should therefore be interpreted as exploratory rather than definitive.

Notably, reverse-direction analysis suggested that BMI also had a strong causal effect on OSA, indicating that the relationship between adiposity and OSA may be bidirectional. This pattern is biologically plausible because obesity may promote upper airway collapse^24^, whereas OSA-related intermittent hypoxia and neuroendocrine dysregulation may further aggravate metabolic dysfunction^25^. Taken together, these findings suggest that BMI may function not only as a partial mediator, but also as an upstream susceptibility factor within a broader reciprocal relationship linking OSA and cardiometabolic risk. This also implies that the role of adiposity in the OSA–MI association may be partly overlapping and not fully captured by a simple linear mediation framework.

AF showed a clearer downstream pattern than BMI. The causal signal from OSA to AF was stronger than the reverse-direction association, which is consistent with previous clinical observations that OSA may contribute to atrial remodeling and arrhythmogenesis^26,27^. In the present study, this directional pattern supports the possibility that AF may represent an important downstream component in the transmission of OSA-related cardiovascular risk. AF may therefore reflect not only a potential mediator, but also a marker of downstream electrical and structural cardiac remodeling related to OSA^28^. However, the precise biological and clinical mechanisms linking OSA, AF, and MI remain to be clarified, and the extent to which AF directly mediates MI risk, rather than reflecting broader cardiovascular remodeling or shared susceptibility, remains uncertain.

SBP is widely recognized as an important downstream phenotype of OSA in clinical practice^29^, but in our mediation analyses it did not show the expected dominant role. Although SBP was strongly associated with MI in multivariable models, its estimated mediation proportion in the OSA–MI pathway was minimal. One possible explanation is that SBP may represent a parallel downstream consequence or correlated component of cardiovascular risk, rather than the principal mediator of the OSA effect. Another possibility is that currently available blood pressure genetic instruments, which are largely derived from clinic-based resting blood pressure measurements, may not adequately capture nocturnal or time-specific hemodynamic abnormalities related to OSA^30,31^. In particular, OSA-related hemodynamic burden may be expressed more through nocturnal blood pressure surges, non-dipping patterns, and short-term variability than through average clinic-based SBP. Importantly, the limited mediation signal for SBP should not be interpreted as evidence that blood pressure is unimportant in OSA-related cardiovascular disease. Rather, it suggests that the role of blood pressure in this pathway may be more complex than that suggested by conventional mediation models.

The progressive multivariable Mendelian randomization analyses further refined the interpretation of obesity-related pathways. After simultaneous modeling of BMI and waist-to-hip ratio (WHR), the apparent independent effect of BMI was attenuated, whereas WHR emerged as a more important factor. This finding suggests that central adiposity may be more closely related to the transmission of cardiovascular risk than overall adiposity^32^. However, this pattern may reflect not only biological differences between overall and central adiposity, but also the difficulty of separating closely correlated adiposity traits within multivariable models^33^. At the same time, the disappearance of the BMI signal in multivariable models should not be interpreted as evidence that general obesity is clinically unimportant. Rather, it suggests that WHR may capture the more proximal component of adiposity-related risk in this specific pathway. Accordingly, the relative prominence of WHR in this analysis should be interpreted as pathway-specific rather than as evidence that BMI is generally less clinically relevant.

After adjustment for type 2 diabetes and lipid-related traits, OSA still retained a significant association with MI that was independent of the metabolic traits included in the model. This finding suggests that the cardiovascular effect of OSA may not be fully explained by conventional metabolic pathways alone. However, this does not imply that metabolic dysfunction is unimportant, but rather that the metabolic traits included here may not fully capture the biological pathways through which OSA contributes to cardiovascular risk. A possible explanation is that OSA-related chronic intermittent hypoxia may contribute directly to cardiovascular injury through mechanisms such as sympathetic activation, oxidative stress, endothelial dysfunction, or myocardial stress^34,35^. These mechanisms provide a biologically plausible interpretation of the residual association, although they were not directly tested in the present study and require further investigation.

When AF and smoking were additionally incorporated into the multivariable models, the direct effect of OSA was further attenuated. In the head-to-head model including both AF and SBP, AF remained independently associated with MI despite the strong effect of SBP, whereas the direct effect of OSA was only marginally significant. These findings suggest that the cardiovascular consequences of OSA may not be fully explained by hemodynamic load alone and support the possibility that AF represents an important downstream component of the OSA–MI pathway. In this context, AF may capture aspects of downstream arrhythmic or remodeling-related risk that are not fully represented by SBP alone. However, the persistence of the AF signal in multivariable models should not be taken to indicate that AF is necessarily a direct mediator in all cases, as it may also reflect broader electrical and structural cardiac remodeling^36^. Even so, the residual borderline effect of OSA after adjustment suggests that additional mediating or direct mechanisms may also exist.

Taken together, the present findings have potential clinical implications. They suggest that cardiovascular risk assessment in OSA may need to extend beyond relief of upper airway obstruction alone. Greater attention may need to be paid to adiposity, especially central adiposity, as well as to arrhythmic phenotypes such as AF, which may represent clinically relevant components of downstream cardiovascular risk. However, these implications should be interpreted cautiously, because Mendelian randomization primarily informs disease etiology rather than the expected benefit of specific interventions. Rather, the present findings may help identify clinical domains that warrant closer attention in cardiovascular risk stratification among patients with OSA. More integrated, multidimensional approaches may therefore better reflect the complexity of OSA-related cardiovascular risk, particularly in high-risk individuals.

Our study has several strengths. First, we used a Mendelian randomization framework with large-scale GWAS data to investigate the association between OSA and MI from a causal inference perspective. Second, we incorporated multiple sensitivity analyses and external validation analyses, which reduced reliance on any single analytical assumption and strengthened the overall robustness of the findings. Third, by combining 2-step Mendelian randomization with progressive multivariable Mendelian randomization, we were able to move beyond a simple exposure–outcome association and examine potential intermediate pathways in a more structured and hypothesis-generating manner.

This study also has several limitations. First, because currently available GWAS data largely define OSA as a binary phenotype, we were unable to model the full spectrum of nocturnal hypoxic burden, sleep-related physiological disturbance, or disease severity. Accordingly, the genetic instruments used here are more likely to capture overall liability to OSA than specific pathophysiological dimensions of the disorder. Second, the instrument selection threshold for OSA was less stringent than the conventional genome-wide significance level. Although instrument strength appeared acceptable, this approach may still have increased the likelihood of including variants with limited specificity to OSA. Third, the study population was primarily of European ancestry, which may limit the generalizability of the findings to other populations. Fourth, some mediation results, particularly for BMI, did not remain significant after multiple-testing correction and should therefore be regarded as exploratory rather than definitive. In addition, because several candidate mediators were closely correlated, the multivariable Mendelian randomization analyses may have been affected by difficulty in disentangling their independent effects^37^. Finally, although our analyses suggest several potential pathways linking OSA and MI, they identify pathway-level signals rather than directly measured biological mechanisms, which remain incompletely understood and warrant further study.

In conclusion, our findings support evidence for a causal association between OSA and MI and suggest that this relationship may be transmitted through a multidimensional network involving adiposity, arrhythmic remodeling, and other cardiometabolic processes. In particular, AF may represent a downstream component of risk that is not fully explained by blood pressure-related load. Further studies are needed to clarify these mechanisms and to determine how these findings may help inform future cardiovascular risk stratification and prevention in patients with OSA.

## Data Availability

The summary-level data used in this study were obtained from publicly available genome-wide association study datasets, including FinnGen, UK Biobank, CARDIoGRAM, GIANT, DIAGRAM, MAGIC, GLGC, CKDGen, GUGC, and GSCAN. Detailed information on all data sources and GWAS IDs is provided in Supplementary Table 1. These datasets are available from the corresponding consortia or through publicly accessible resources such as the IEU OpenGWAS platform.

https://www.diagram-consortium.org/

https://giant-consortium.web.broadinstitute.org/

https://cardiogramplusc4d.org/

https://www.ukbiobank.ac.uk/

https://www.finngen.fi/en/access_results

## Acknowledgements

We thank the participants and investigators of FinnGen, UK Biobank, CARDIoGRAM, GIANT, DIAGRAM, MAGIC, GLGC, CKDGen, GUGC, and GSCAN for making their GWAS summary statistics publicly available. We also acknowledge the IEU OpenGWAS/MR-Base platform for data access. Their contributions made this Mendelian randomization study possible.

## Supplementary Materials

**Supplementary Table 1.**
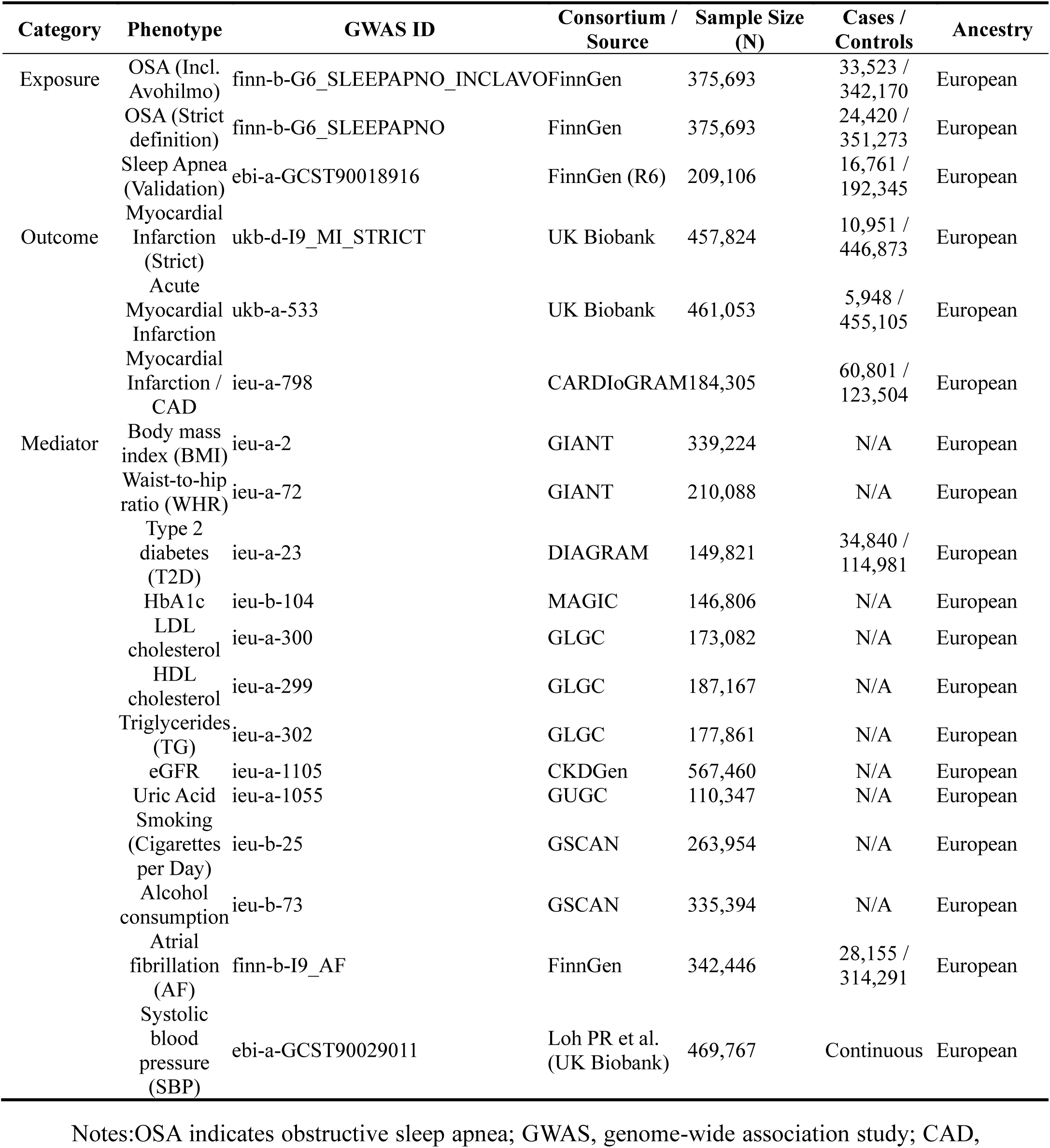
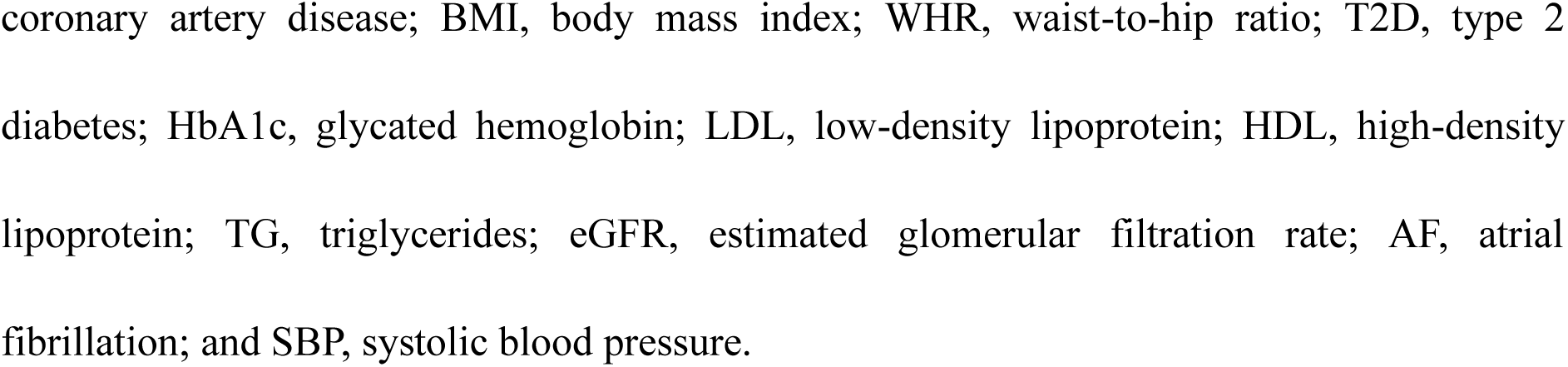
Details of GWAS data sources used for OSA, myocardial infarction, and potential mediators.

**Supplementary Table 2.** Characteristics of the instrumental variables for obstructive sleep apnea used in the primary Mendelian randomization analysis.

| rsID | Chromosome | Position | Effect Allele | Other Allele | EAF (Exposure) | Beta (Exposure) | SE (Exposure) | P-value (Exposure) |
| --- | --- | --- | --- | --- | --- | --- | --- | --- |
| rs1023230 | 13 | 20270158 | T | C | 0.9223 | -0.1116 | 0.0232 | 1.52398e-0 |
| rs10507084 | 12 | 97753152 | T | C | 0.1794 | 0.1076 | 0.0163 | 4.06912e-1 |
| rs10512749 | 5 | 4839617 | T | G | 0.0222 | 0.1874 | 0.0423 | 9.58098e-0 |
| rs10860169 | 12 | 97671568 | G | A | 0.2906 | -0.0645 | 0.0137 | 2.68701e-0 |
| rs10910079 | 1 | 2394648 | T | C | 0.03324 | 0.157 | 0.0348 | 6.25302e-0 |
| rs10928560 | 2 | 136991807 | T | C | 0.1948 | -0.0874 | 0.0158 | 3.17497e-0 |
| rs10938398 | 4 | 45186139 | A | G | 0.4731 | 0.058 | 0.0125 | 3.145e-06 |
| rs113241098 | 6 | 24896688 | G | A | 0.0241 | 0.1816 | 0.0404 | 7.01795e-0 |
| rs11530654 | 11 | 100364301 | C | A | 0.2633 | 0.0659 | 0.0141 | 3.05999e-0 |
| rs116497662 | 3 | 17996736 | A | C | 0.05604 | 0.1203 | 0.0272 | 9.89305e-0 |
| rs116528678 | 10 | 57381308 | A | G | 0.002934 | -0.5443 | 0.1224 | 8.73092e-0 |
| rs11758441 | 6 | 43813586 | T | C | 0.378 | 0.0596 | 0.0129 | 3.61302e-0 |
| rs117670722 | 20 | 13786428 | T | A | 0.06119 | -0.1172 | 0.0263 | 8.31304e-0 |
| rs12205015 | 6 | 113984567 | T | A | 0.1125 | -0.0899 | 0.0198 | 5.49098e-0 |
| rs1223589 | 10 | 95759405 | A | G | 0.2159 | -0.0686 | 0.0151 | 5.90704e-0 |
| rs12682930 | 9 | 110379567 | T | G | 0.06093 | -0.128 | 0.0262 | 1.064e-06 |
| rs12890119 | 14 | 29068403 | T | C | 0.451 | 0.0551 | 0.0125 | 9.64295e-0 |
| rs1378892 | 15 | 51964865 | C | T | 0.8563 | -0.0798 | 0.0177 | 6.35097e-0 |
| rs1403763 | 3 | 135796240 | G | C | 0.2283 | 0.0679 | 0.0149 | 5.17202e-0 |
| rs142006783 | 16 | 74651733 | C | T | 0.03779 | 0.1781 | 0.0327 | 4.95405e-0 |
| rs1425898 | 8 | 54115304 | G | A | 0.6755 | 0.0618 | 0.0139 | 8.72208e-0 |
| rs1454933 | 12 | 33017043 | C | A | 0.06345 | 0.1152 | 0.0256 | 6.50894e-0 |
| rs149210528 | 14 | 62269750 | C | T | 0.01621 | -0.2233 | 0.0501 | 8.29698e-0 |
| rs149473233 | 3 | 6221580 | A | G | 0.005352 | 0.3975 | 0.0895 | 9.01301e-0 |
| rs181190675 | 3 | 88754727 | T | C | 0.01816 | -0.2223 | 0.0475 | 2.87998e-0 |
| rs182846984 | 4 | 185546497 | T | G | 0.01413 | 0.269 | 0.0544 | 7.55701e-0 |
| rs1896039 | 12 | 55963016 | A | G | 0.5349 | 0.0624 | 0.0126 | 7.55005e-0 |
| rs193546 | 14 | 72660200 | A | G | 0.7438 | 0.0731 | 0.0143 | 3.02803e-0 |
| rs1959185 | 14 | 84112949 | A | G | 0.1344 | 0.0844 | 0.0182 | 3.51998e-0 |
| rs2725231 | 4 | 88934955 | G | A | 0.8335 | -0.0755 | 0.0166 | 5.51201e-0 |
| rs3996329 | 7 | 2018870 | A | G | 0.2366 | -0.0768 | 0.0148 | 1.98701e-0 |
| rs405430 | 9 | 101473404 | T | G | 0.7624 | -0.0696 | 0.0146 | 1.88101e-0 |
| rs4244535 | 11 | 28457737 | T | C | 0.499 | 0.0552 | 0.0124 | 9.02506e-0 |
| rs4837016 | 9 | 128141809 | A | G | 0.4662 | -0.0697 | 0.0125 | 2.29699e-0 |
| rs4934470 | 10 | 91067438 | C | T | 0.7976 | 0.0693 | 0.0155 | 7.93597e-0 |
| rs4961731 | 9 | 16683543 | T | C | 0.8895 | 0.0962 | 0.02 | 1.52901e-0 |
| rs527014 | 1 | 53977292 | T | C | 0.07431 | 0.1231 | 0.0237 | 2.16202e-0 |
| rs6021831 | 20 | 50890662 | G | C | 0.2974 | -0.0636 | 0.0136 | 2.93103e-0 |
| rs61819581 | 1 | 209253452 | C | G | 0.02472 | 0.1817 | 0.04 | 5.63599e-0 |
| rs61842991 | 10 | 14805937 | C | T | 0.2607 | -0.0636 | 0.0142 | 7.67803e-0 |
| rs62385672 | 5 | 168027543 | C | G | 0.02366 | -0.1862 | 0.0415 | 7.13493e-0 |
| rs6791437 | 3 | 168616977 | G | A | 0.4097 | -0.0569 | 0.0127 | 7.43995e-0 |
| rs6845679 | 4 | 92350342 | T | C | 0.5898 | 0.0582 | 0.0127 | 4.28401e-0 |
| rs72632980 | 11 | 16653971 | C | T | 0.07216 | 0.1167 | 0.0241 | 1.24601e-0 |
| rs72892016 | 18 | 38872987 | A | G | 0.1204 | -0.0886 | 0.0192 | 3.804e-06 |
| rs72953990 | 1 | 88064857 | A | G | 0.1932 | -0.071 | 0.0159 | 7.75193e-0 |
| rs7312617 | 12 | 102931304 | G | A | 0.7157 | -0.0633 | 0.0138 | 4.54601e-0 |
| rs770267 | 13 | 33691082 | G | A | 0.831 | 0.0767 | 0.0166 | 4.06303e-0 |
| rs77998028 | 3 | 146780513 | T | C | 0.00632 | 0.3524 | 0.0794 | 9.13692e-0 |
| rs78730556 | 4 | 125311740 | T | C | 0.06739 | -0.1135 | 0.0251 | 5.87801e-0 |
| rs80079566 | 19 | 57676371 | C | T | 0.01355 | 0.2447 | 0.0551 | 9.03109e-0 |
| rs9510253 | 13 | 23181364 | T | A | 0.1468 | 0.0826 | 0.0176 | 2.666e-06 |
| rs9937053 | 16 | 53799507 | A | G | 0.4299 | 0.1021 | 0.0125 | 4.02717e-1 |
| rs996762 | 2 | 59772059 | G | C | 0.8239 | -0.0804 | 0.0163 | 7.90296e-0 |

Notes: rsID indicates reference SNP identifier; EAF, effect allele frequency; SE, standard error. Beta (Exposure) and Beta (Outcome) represent the SNP effect estimates for obstructive sleep apnea and myocardial infarction, respectively. R-squared indicates the proportion of variance in obstructive sleep apnea explained by each instrument.

**Supplementary Table 3.** List of 21 genetic variants excluded due to potential pleiotropic associations with cardiovascular and metabolic confounders.

| SNP (rsID) | Associated Phenotypes (GWAS Traits) | Primary Reasoning for Exclusion |
| --- | --- | --- |
| rs6845679 | Smoking initiation, Smoking status | Pleiotropy via tobacco use |
| rs10475978 | Smoking initiation, Waist circumference<br>adj. BMI | Pleiotropy via smoking and abdominal adiposity |
| rs1403763 | Type 2 diabetes, BMI, Waist circumference<br>adj. BMI | Pleiotropy via glucose and general adiposity |
| rs12205015 | Body mass index (BMI) | Pleiotropy via general obesity |
| rs2725231 | Urate levels | Pleiotropy via urate metabolism |
| rs1223589 | Waist-hip ratio (WHR) | Pleiotropy via body fat distribution |
| rs1378892 | BMI, WHR, Type 2 diabetes | Multi-system pleiotropy (Obesity & Diabetes) |
| rs4244535 | Serum urate levels, Childhood BMI | Pleiotropy via urate and early-life adiposity |
| rs9937053 | BMI, Waist circumference, Type 2 diabetes | Pleiotropy via metabolic syndrome components |
| rs4837016 | Smoking (status, cessation, initiation) | Comprehensive pleiotropy via tobacco behavior |
| rs10928560 | Urate levels, Body mass index | Pleiotropy via urate and obesity |
| rs3996329 | Smoking initiation, BMI, Type 2 diabetes | Multi-pathway metabolic and lifestyle pleiotropy |
| rs996762 | Smoking initiation | Pleiotropy via tobacco use |
| rs72632980 | Body mass index | Pleiotropy via general obesity |
| rs4961731 | Urate levels | Pleiotropy via urate metabolism |
| rs405430 | Body mass index | Pleiotropy via general obesity |
| rs10860169 | Age at smoking initiation | Pleiotropy via lifestyle factor (smoking) |
| rs6021831 | BMI, Waist circumference, Type 2 diabetes | Pleiotropy via metabolic syndrome components |
| rs10938398 | BMI, Waist circumference, Type 2 diabetes | Pleiotropy via metabolic syndrome components |
| rs11758441 | WHR adj. BMI, Urate levels, Type 2 diabetes | Multi-pathway metabolic pleiotropy |
| rs72892016 | Smoking initiation | Pleiotropy via tobacco use |

Notes: SNP indicates single-nucleotide polymorphism; rsID, reference SNP identifier;
GWAS, genome-wide association study; BMI, body mass index; and WHR, waist-to-hip ratio.
Variants were excluded when previously reported trait associations indicated possible horizontal pleiotropy through cardiovascular, metabolic, or lifestyle-related pathways.

**Supplementary Table 4.** Characteristics of the 9 instrumental variables used in the reverse Mendelian randomization analysis (myocardial infarction on obstructive sleep apnea).

| rsID | Chromosome | Position | Effect Allele | Other Allele | EAF (Exposure) | Beta (Exposure) | SE (Exposure) | P-value (Exposure) |
| --- | --- | --- | --- | --- | --- | --- | --- | --- |
| rs10455872 | 6 | 161010118 | G | A | 0.0807558 | 0.0065218 | 0.000591863 | 3.123 |
| rs12740374 | 1 | 109817590 | T | G | 0.221809 | -0.00226118 | 0.000387831 | 5.537 |
| rs1333042 | 9 | 22103813 | G | A | 0.498058 | 0.00398365 | 0.000322534 | 4.883 |
| rs1510226 | 6 | 160816409 | C | T | 0.0195761 | 0.00945841 | 0.00116823 | 5.681 |
| rs1909196 | 1 | 222786032 | C | T | 0.679605 | 0.00189512 | 0.000346033 | 4.335 |
| rs28451064 | 21 | 35593827 | A | G | 0.13177 | 0.00279752 | 0.000486771 | 9.086 |
| rs35350651 | 12 | 111907431 | AC | A | 0.503996 | -0.0023299 | 0.000322568 | 5.096 |
| rs814573 | 19 | 45424351 | T | A | 0.187345 | 0.00269299 | 0.000424053 | 2.147 |
| rs9349379 | 6 | 12903957 | G | A | 0.405522 | 0.00202898 | 0.000328294 | 6.401 |

Notes: Abbreviations: SNP, single-nucleotide polymorphism; Chr, chromosome; Pos, position; EAF, effect allele frequency; SE, standard error; $R^2$ , proportion of variance explained; F, F-statistic. The exposure dataset is myocardial infarction from the UK Biobank, and the outcome dataset is obstructive sleep apnea from FinnGen.

**Supplementary Figure 1.**
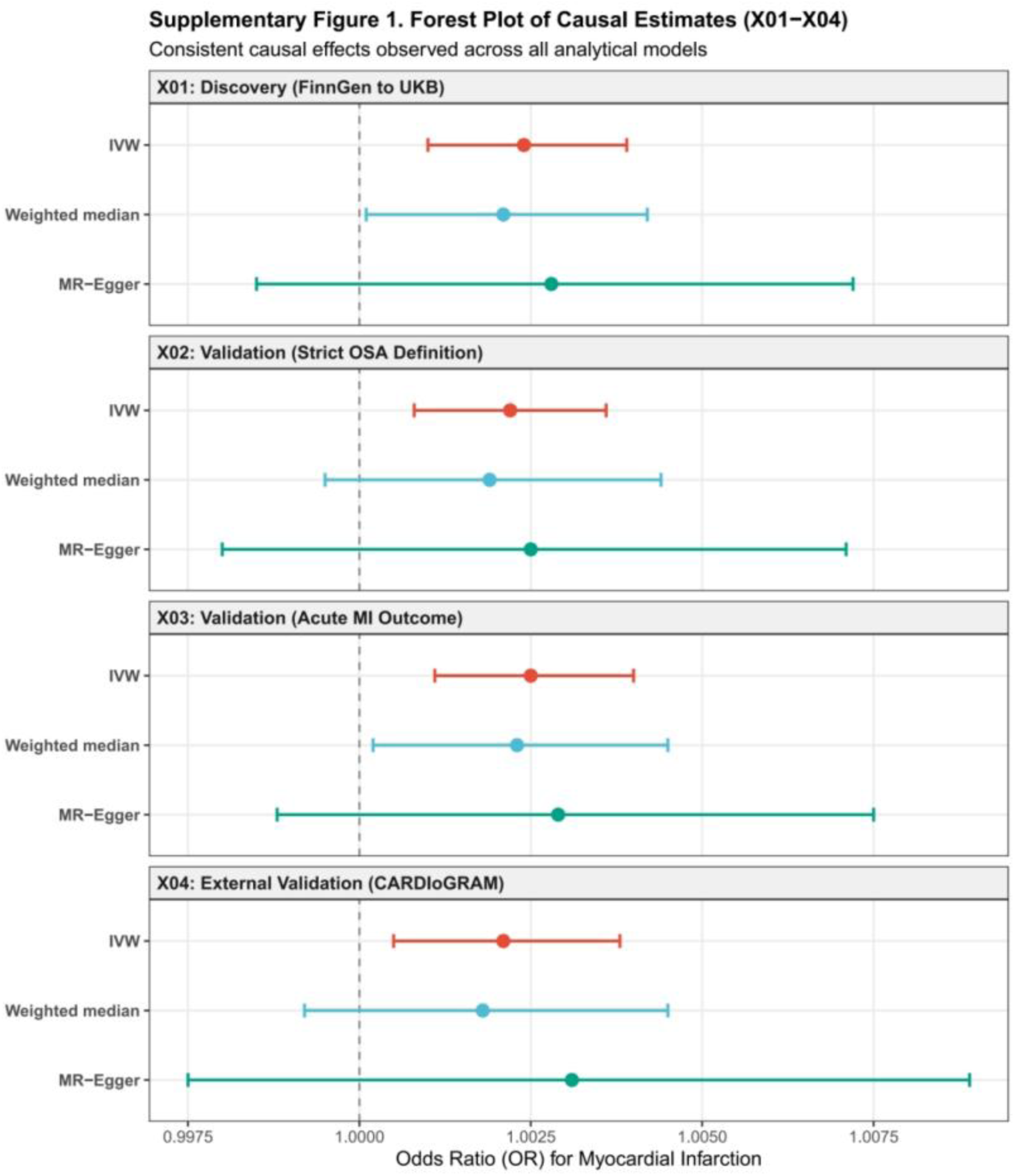
Forest plot of Mendelian randomization causal estimates across discovery and validation models. The plot summarizes the causal odds ratios (ORs) and 95% confidence intervals (CIs) for the effect of obstructive sleep apnea (OSA) on myocardial infarction (MI) using four different analytical models: X01 (primary discovery), X02 (strict OSA definition), X03 (acute MI outcome), and X04 (external validation using the CARDIoGRAM consortium). Within each model, three MR methods were applied: inverse-variance weighted (IVW), weighted median, and MR-Egger regression. The consistent results across all models and methods underscore the robustness of the positive causal association between OSA and MI risk.

**Supplementary Figure 2.**
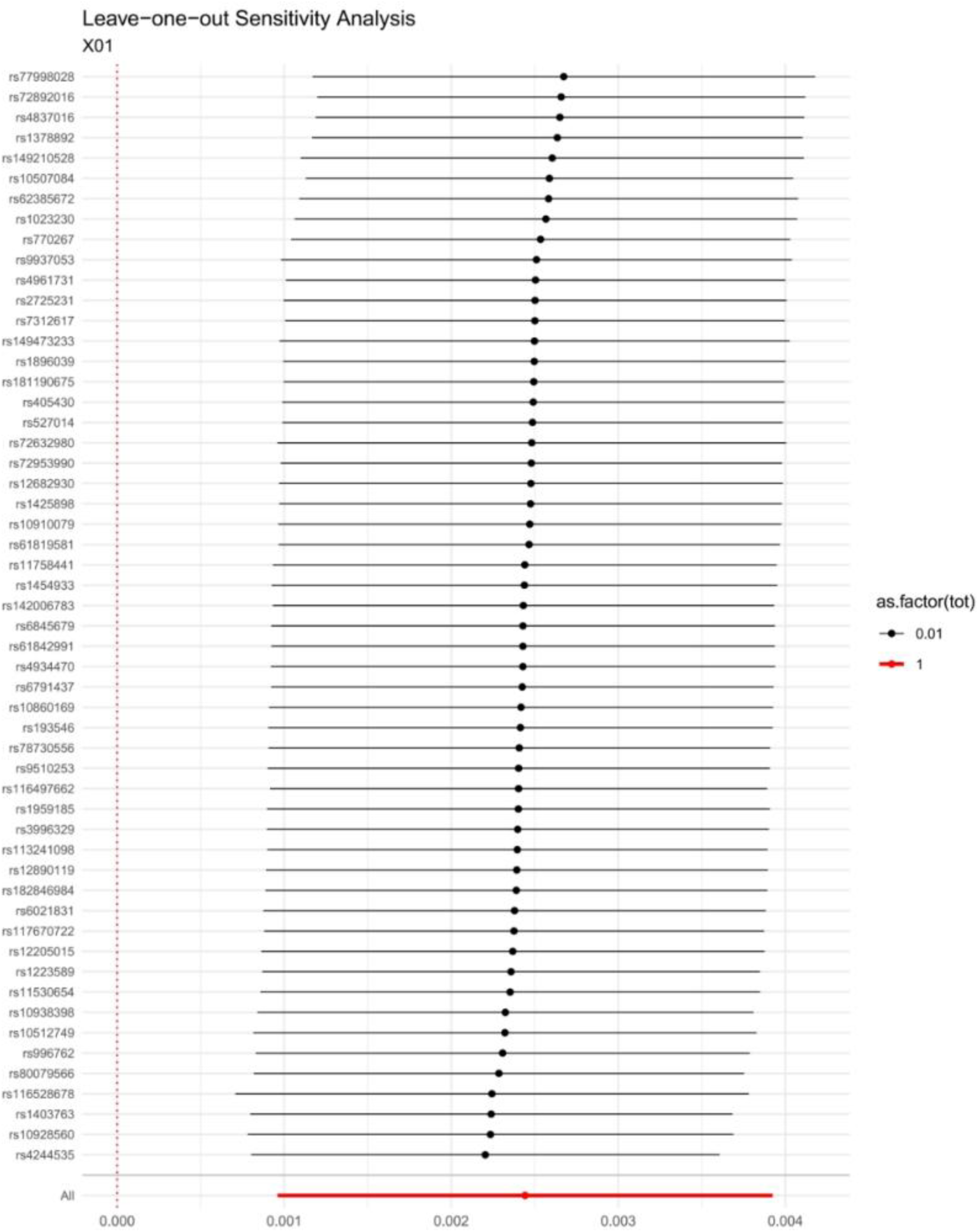
Leave-one-out sensitivity analysis for the causal effect of OSA on MI (X01). Each horizontal line represents the MR instrumental variable weighted (IVW) estimate and 95% confidence interval for the effect of obstructive sleep apnea (OSA) on myocardial infarction (MI) after excluding that specific single nucleotide polymorphism (SNP) from the analysis. The red point at the bottom represents the IVW estimate using all instrumental variables. The fact that all confidence intervals remain consistently above zero, regardless of which SNP is excluded, demonstrates that the overall causal association is robust and not driven by any single outlier variant.

**Supplementary Figure 3.**
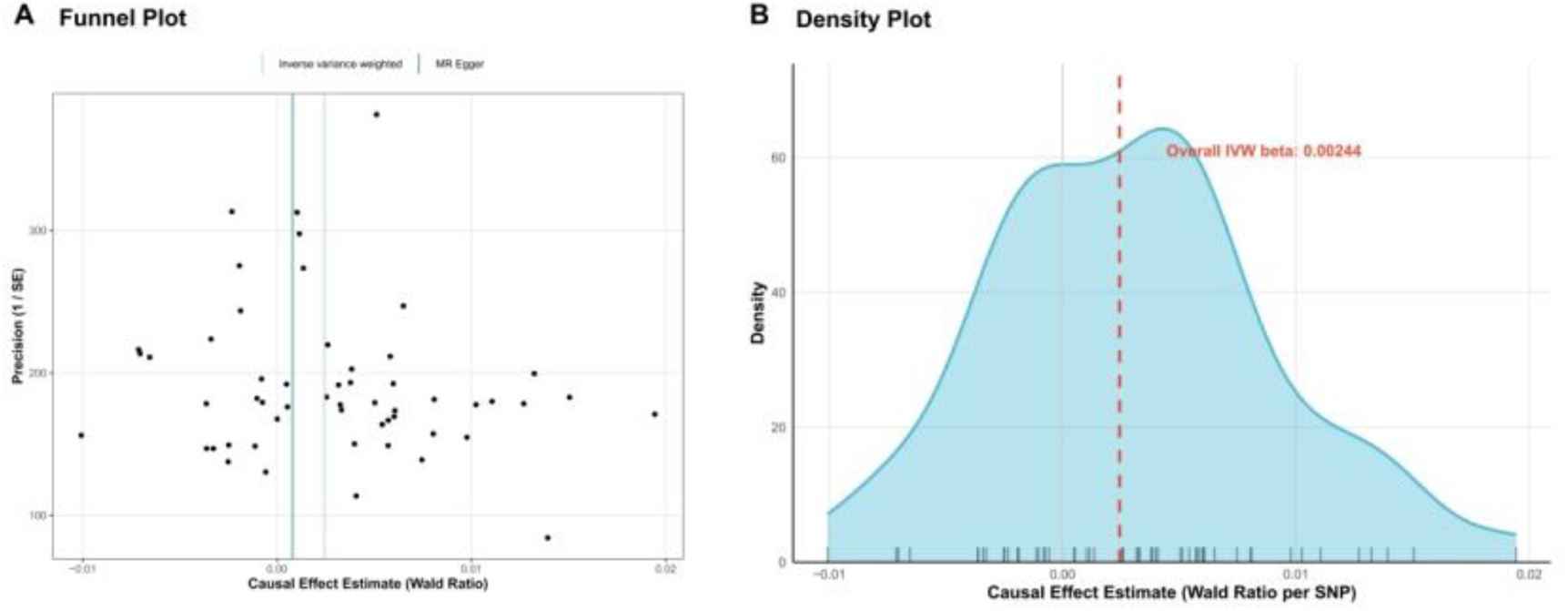
Diagnostic plots for the causal effect of OSA on MI in the discovery model (X01). (A) Funnel plot showing the relation between the causal effect of each individual SNP (Wald ratio) and its precision (1/SE). The symmetrical distribution around the IVW and MR-Egger lines indicates the absence of significant directional pleiotropy. (B) Density plot illustrating the distribution of individual SNP causal estimates. The single prominent peak aligned with the overall IVW estimate (red dashed line) suggests highly consistent causal signals across the instrumental variables.

**Supplementary Figure 4.**
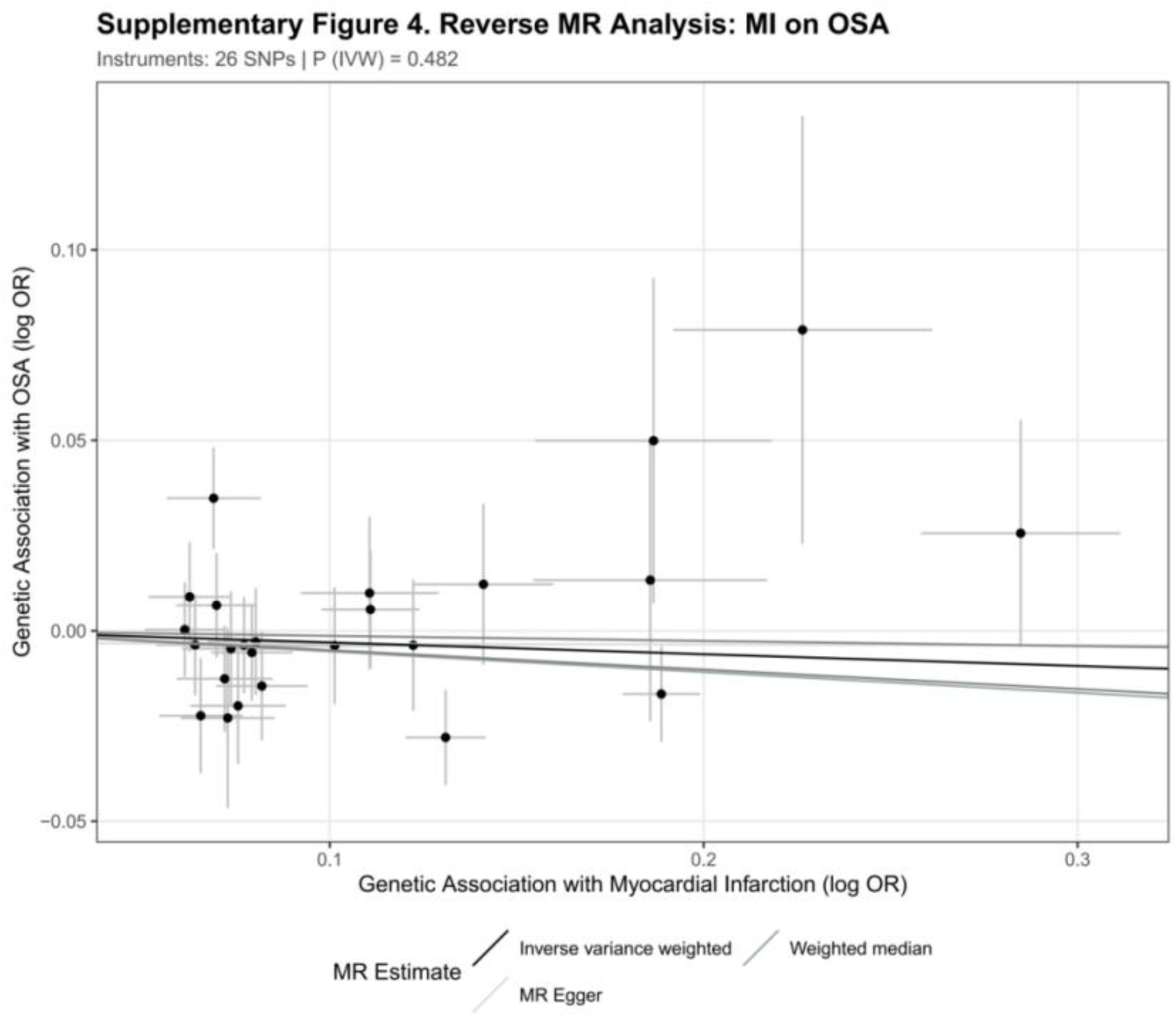
Scatter plot of the reverse Mendelian randomization analysis examining the causal effect of MI on OSA risk. The scatter plot displays the associations of genetic variants with myocardial infarction (MI, exposure) against their associations with obstructive sleep apnea (OSA, outcome). Each point represents a genetic instrument for MI. The regression lines represent different MR methods (IVW, MR-Egger, and weighted median). The horizontal orientation of the fitting lines and the non-significant P-value P=0.482 indicate no evidence of reverse causality, confirming that MI does not causally influence the risk of OSA. Distinct gray tones, including black for IVW, are used to represent each MR method while maintaining a neutral palette suitable for presenting null findings.

